# Evidence for antibody as a protective correlate for COVID-19 vaccines

**DOI:** 10.1101/2021.03.17.20200246

**Authors:** Kristen A. Earle, Donna M. Ambrosino, Andrew Fiore-Gartland, David Goldblatt, Peter B. Gilbert, George R. Siber, Peter Dull, Stanley A. Plotkin

**Affiliations:** Bill & Melinda Gates Foundation; Independent Advisor; Fred Hutchinson Cancer Research Center; University College London; University of Pennsylvania

## Abstract

Though eleven novel COVID-19 vaccines have demonstrated efficacy, additional affordable, scalable, and deliverable vaccines are needed to meet unprecedented global demand. With placebo-controlled efficacy trials becoming infeasible due to the roll out of licensed vaccines, a correlate of protection is urgently needed to provide a path for regulatory approval of novel vaccines. To assess whether antibody titers may reasonably predict efficacy, we evaluated the relationship between efficacy and in vitro neutralizing and binding antibodies of 7 vaccines for which sufficient data have been generated. Once calibrated to titers of human convalescent sera reported in each study, a robust correlation was seen between neutralizing titer and efficacy (ρ= 0.79) and binding antibody titer and efficacy (ρ = 0.93), despite geographically diverse study populations subject to different forces of infection and circulating variants, and use of different endpoints, assays, convalescent sera panels and manufacturing platforms. This correlation is strengthened by substituting post-hoc analyses for efficacy against the ancestral strain (D614G), where available. Together with an accumulating body of evidence from natural history studies and animal models, these results support the use of post-immunization antibody titers as the basis for establishing a correlate of protection for COVID-19 vaccines.

## Full Text

Eleven novel COVID-19 vaccines have demonstrated efficacy with several more undergoing Phase 3 clinical trials. Despite this, to meet unprecedented global demand, additional vaccines are needed even as placebo-controlled efficacy trials are becoming infeasible. A correlate of protection (CoP) is urgently needed to provide a path for regulatory approval of new vaccines.

Correlates of protection based on antibody levels or functional activity derived from efficacy trials have been used to license many vaccines^1^. We therefore assessed the relationship between efficacy and both virus neutralizing antibody (VNA) and Spike protein-binding IgG antibody for the seven COVID-19 vaccines with sufficient data: Pfizer, Moderna, Gamaleya, AstraZeneca, Sinovac, Novavax, and Janssen. We first evaluated peak geometric mean titers (GMT) of VNA and binding antibodies 1-4 weeks following the recommended vaccination regimen as reported by each manufacturer but found low correlations with efficacy (Figure S1), most likely because assays were not calibrated to a common standard. We then calibrated assays against an imperfect but “best available” standard, titers of human convalescent serum (HCS) reported in each study, revealing a high correlation between neutralizing titer and efficacy (ρ= 0.79) and binding antibody titer and efficacy (ρ = 0.93). In this analysis, neutralizing or IgG binding antibody accounted for 77.5% and 94.2%, respectively, of the variation in efficacy observed among the seven vaccines. A robust correlation was seen despite geographically diverse study populations which were subject to different forces of infection and circulating variants, and use of different endpoints, assays and manufacturing platforms (Figure 1, Tables S1-S2; additional analyses in Figures S1-S4). Because correlation does not prove causation, other factors in addition to antibody may impact efficacy.

**Figure 1.**
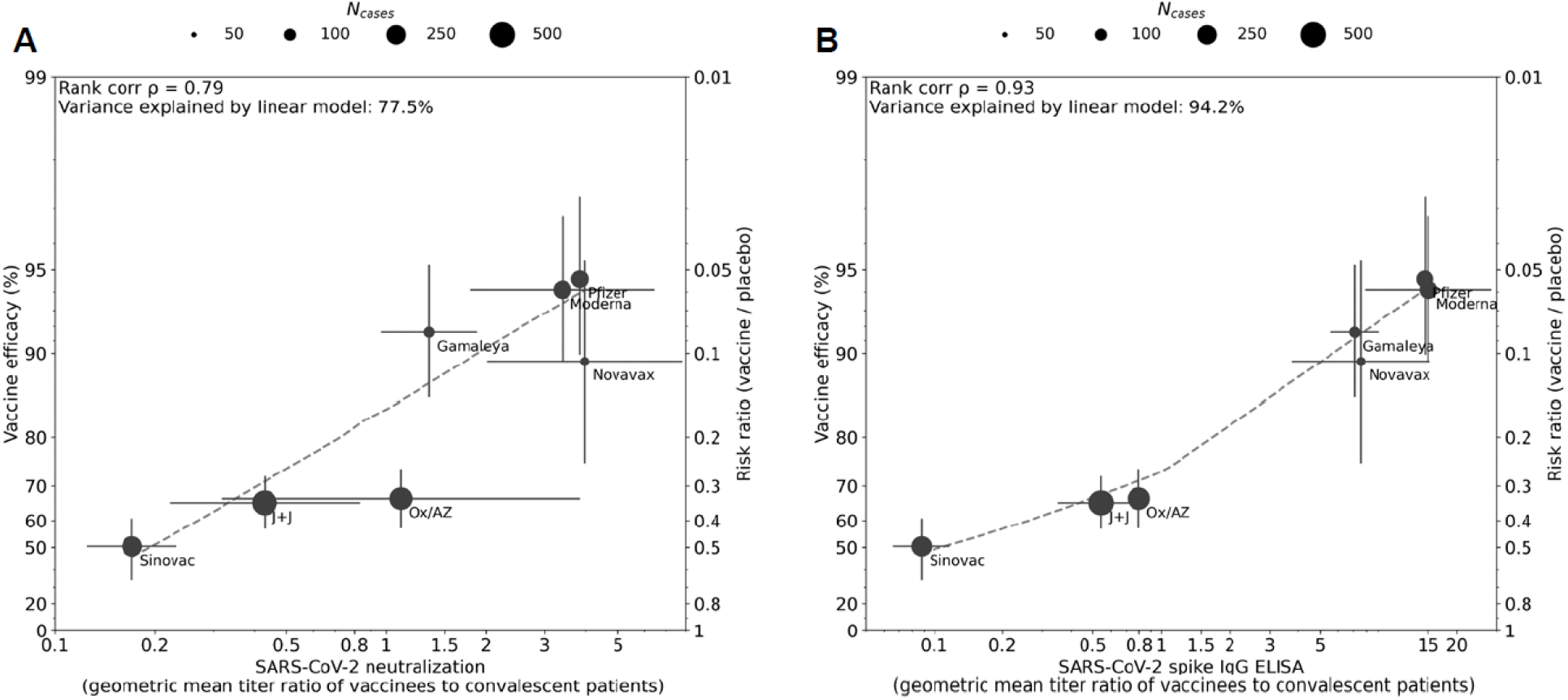
Correlation between antibody responses and efficacy rate for 7 COVID-19 vaccines. Panels A and B display correlations of antibody responses for neutralization and ELISA assay ratios, respectively. Data included in correlation analyses, including efficacy, vaccine-induced and human convalescent sera (HCS) geometric mean antibody responses, description of HCS panels, and ratios calibrating vaccine-induced responses to HCS are described in Tables S1 and S2. Where multiple efficacy estimates exist for a single product, the primary planned analysis and endpoints from Phase 3 studies were selected. Neutralization antibody geometric mean titers were generated from wild-type virus neutralization assays for all vaccines except Moderna and AstraZeneca / Oxford, where pseudoviral-based assays were utilized. Anti-SARS-CoV-2 binding titers were generated by anti-Spike protein ELISAs, except for Moderna, Gamaleya, and Sinovac, which utilized an anti-Receptor Binding Domain ELISA, and Pfizer/BioNTech which utilized an anti-S1 ELISA. Dot size corresponds to the number of cases reported for Phase 3 efficacy analyses. The y-axis is estimated log risk ratio reported on the vaccine efficacy scale. The x-axis is ratio of the peak geometric mean neutralization titer or ELISA titer at 7-28 days post vaccination, relative to HCS. Error bars indicate 95% Confidence Intervals (except for Oxford/AZ antibody responses, which represent ratios of median titers with interquartile ranges) with dashed line showing non-parametric LOESS fit. A rank correlation value was calculated with r^2^ in a linear model utilized for variance explanation.

The results support the use of post-immunization antibody levels as the basis for a CoP. We propose that the next steps toward achieving consensus on a CoP include the following: First, to establish comparability of antibody measurements among laboratories by (i) using the WHO International Standard (NIBSC 20/136) to express VNA titers in IU/ml and binding antibody titers in BAU/ml and (ii) establishing a relevant proficiency panel. Second, to agree on an assay, most likely a VNA meeting performance-based criteria, to serve as the gold standard assay for CoP and perhaps to allow validation of secondary assays by strong correlations with the gold standard. Third, to calculate the protective threshold in each phase 3 study based either on the distribution of antibody titers in random samples of vaccinees and controls^2^ or on case-cohort evaluations^3^. Fourth, since the CoP calculated from different studies may differ, stakeholders should be convened to arrive at a consensus on a minimum protective antibody level for the primary outcome of symptomatic COVID. If possible, it would also be useful to estimate thresholds for preventing severe disease or asymptomatic infection. Fifth, to verify that the CoP will apply to new variants using appropriate adapted assays as information accrues on the immune response and efficacy of vaccines against them.

A CoP agreed to by regulators will support the demonstration of efficacy of new vaccines using traditional criteria for establishing non-inferiority based on the proportion of vaccinees achieving the protective threshold and comparable GMTs, relative to an approved comparator vaccine. Since the relationship between antibody responses and efficacy was consistent across all manufacturing platforms evaluated to date, even though they may differ in their induction of other antibody functions or T cell responses, an argument can be made that any approved vaccine can serve as a comparator for a future candidate vaccine.

## Data Availability

Data sharing is not applicable to this article as no new data were created or analyzed in this study. All clinical data included in the analysis are in the public domain.

## Notes

### Competing Interest Statement

Dr. Plotkin consults for Janssen and Moderna; Dr. Siber reports personal fees from Clover, other from COVAXX, personal fees from CanSino, personal fees from CureVac, personal fees from Valneva, personal fees and other from Affinivax, outside the submitted work; Dr. Gilbert reports grants and non-financial support from SanofiPasteur, outside the submitted work; Dr. Ambrosino reports personal fees from Covaxx, personal fees from Clover Biopharmaceuticals, outside the submitted work.

### Clinical Trial

N/A

### Funding Statement

The authors received no financial support for the research or authorship of this article.

