## Supplementary material for "Evidence for antibody as a protective correlate for COVID-19 vaccines": Figure S1

Supplementary Appendix

**Table of Contents**

Methods

Immunogenicity data………………………………………………………………………………………………………………………..3

Vaccine efficacy……………………………..……………………………………………..….………………………………………………3

Statistical analysis……………………………………………………………………………………………………………………………..3

Supplementary figures

Figure S1: Direct correlation of antibody titers to efficacy for 7 COVID-19 vaccines…………………………..4

Figure S2: Sensitivity analyses modeling impact of +/- 2-fold changes in HCS values………………………….5

References…………………………………………………………………………………………………….……………………………………….12

Methods

Immunogenicity data

Neutralizing and binding antibody titers were identified from peer-reviewed publications of Phase 1 or Phase 2 clinical studies for inclusion in the primary analysis. Publication of a human convalescent sera (HCS) panel with Phase 1/2 immunogenicity data was required for inclusion in the analysis; where exact values of geometric mean (with 95% confidence intervals) or median (with interquartile range) were not reported, estimates were drawn from the published figure. Where multiple regimens or post-vaccination timepoints were published, the data points that correspond to peak response in the 1-4 weeks following vaccination according to the dose and schedule studied in Phase 3 were chosen. In cases where multiple assays were employed to describe immunogenicity, the assay with the most comprehensive HCS panel run alongside subject data was selected.

Additional publicly available regulatory documents and manuscript pre-prints were considered for evaluation in exploratory analyses included in the supplement. Data summary and sources are provided in Tables S1 and S2.

Vaccine efficacy

COVID-19 vaccines were considered for inclusion in the correlation analysis if interim or primary analysis efficacy point estimates were publicly available at the time of submission from at least one comprehensive source (i.e., peer-reviewed publication, regulatory submission, or release of detailed site- or strain-level data within a press release or briefing). Primary endpoint efficacy analyses were employed to conduct the primary correlation analysis for this study. Post hoc efficacy estimates were considered for evaluation in exploratory analyses included in the supplement.

Statistical analyses

Vaccine efficacy was computed as one minus risk ratio times 100%, and the risk ratio for each study was calculated as specified in the study protocol/primary publication. Rank correlation was the Spearman's rank correlation coefficient (rho) between the readouts on the x- and y-axes across the seven vaccine trials. The percent variance explained in a linear model was computed as the square of the Pearson's correlation coefficient; both x and y data were fit using a natural log transform. No weights were applied to the data to compute correlations or variance explained. The dashed fit line was computed using locally estimated scatterplot smoothing (LOESS) regression (all points fit, with tricube weight function). As a sensitivity analysis we estimated rank correlation coefficients, percent variance explained and linear fit lines from VNA (ELISA) ratios that were simulated using HCS VNA (ELISA) geometric means that were randomly shifted from 0.5-fold to 2-fold from the observed geometric means (uniform sampling over +/- 2-fold range). Datasets were simulated 10K times to estimate the 2.5th and 97.5th percentiles of the rank correlation and percent variance explained; fit lines from 500 simulations were plotted in the figure.

Supplementary correlates analyses were computed using identical methods, but with alternative data points (blue) indicated in the figure legend.

**Figure S1.** **Direct correlation of antibody titers to efficacy for 7 COVID-19 vaccines.** Correlation between vaccine efficacy and un-calibrated geometric mean titers for neutralizing antibodies (Panel A) or binding (full-length Spike, S1, or RBD) IgG antibodies (Panel B) was assessed.

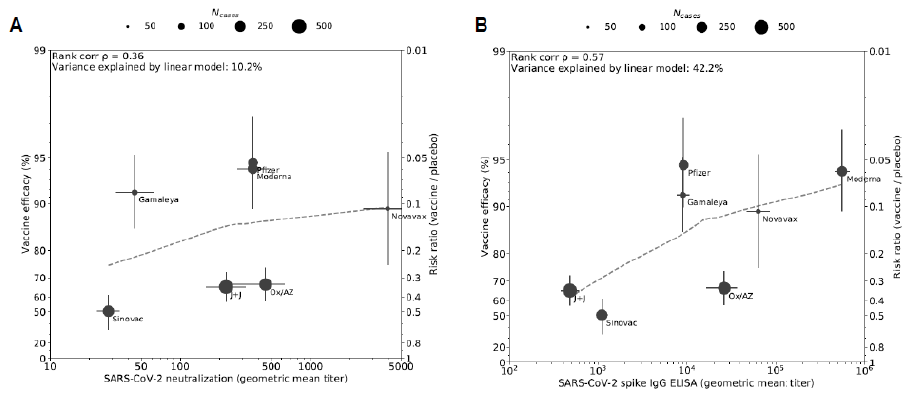

**Figure S2. Sensitivity analyses modeling impact of +/- 2-fold changes in HCS values.** To assess the sensitivity of this analysis to the HCS panel selected by each developer, rank correlation coefficients, percent variance explained and linear fit lines were estimated from VNA (Panel A) and ELISA (Panel B) ratios that were simulated using HCS geometric means that were randomly shifted from 0.5-fold to 2-fold from the observed geometric means (uniform sampling over +/- 2-fold range), consistent with variability seen in HCS across the range of disease severity^1^. Datasets were simulated 10K times to estimate the 2.5th and 97.5th percentiles of the rank correlation and percent variance explained; fit lines from 500 simulations are plotted.

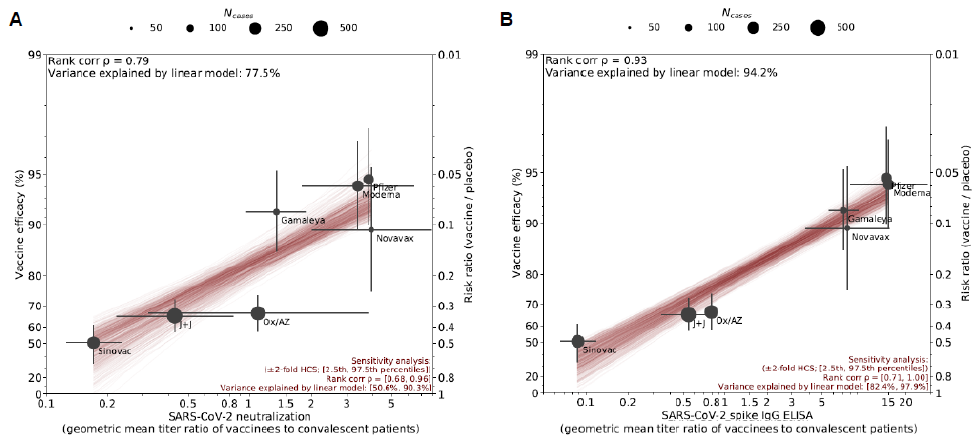

**Figure S3. Impact of post-hoc analyses to assess efficacy against ancestral strain on correlation.** Variability in circulating strains regionally and temporally challenges cross-study comparisons of vaccine efficacy, particularly with the emergence of recent variants of concern (VOCs) that contain Spike protein mutations against which vaccines incorporating ancestral strain Spike antigen may be less effective. To assess the impact that variation in circulating strains may have on correlation between neutralizing antibody titer (Panel A) or binding antibody titer (Panel B) and efficacy, post-hoc analyses that calculate efficacy against the dominant ancestral strain (D614G) or calculate efficacy at sites without circulating VOCs were substituted for primary endpoint efficacy estimates, where available. Post-hoc analyses included are denoted by blue dots. Post-hoc analysis of Novavax vaccine efficacy against the ancestral strain (95.6%) was determined by sequencing 56 of the 62 cases accumulated in the UK Phase 3 study^2^. Vaccine efficacy for the U.S. site of Janssen/J&J’s Phase 3 study (72%) is included based on sequencing of 197 of the 268 cases, suggesting that strain D614G accounted for the vast majority (96.4%) of cases^3^. Binding antibody ratio for J&J (Panel B) was calculated from US-specific Phase 3 data, calibrated to HCS titers published with Phase 1/2 immunogenicity data, as noted in Table S2.

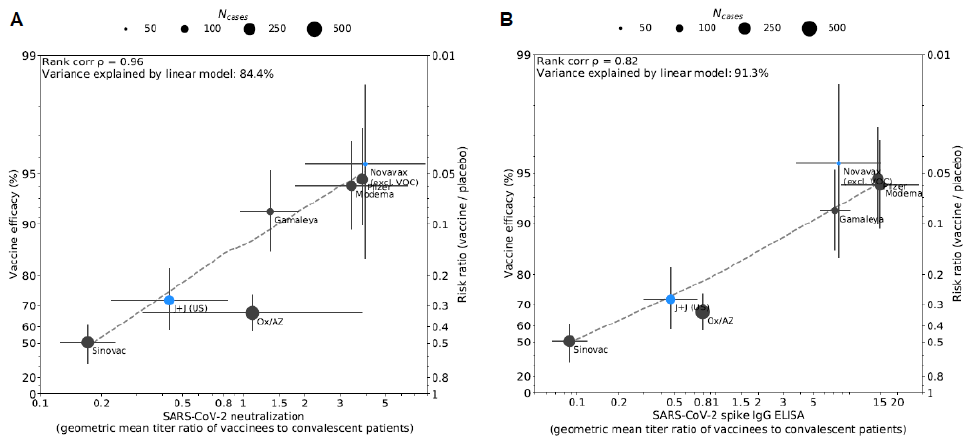

**Figure S4. Impact of exploratory analysis of Oxford/AstraZeneca immunogenicity and efficacy by interval between doses on correlation.** Recent publication of a pooled analysis of Phase 3 trial data for Oxford/AstraZeneca^4^ suggests that interval between doses in Phase 3 studies varied from 4-12+ weeks, and that both vaccine efficacy and immunogenicity data varied by dose interval. The immunogenicity data from Phase 1/2, which corresponded to a 4-week dose interval, may therefore not be representative of immunogenicity generated in the Phase 3 study, or correspond to pooled vaccine efficacy estimates. To assess the impact on correlation, exploratory analyses of dose intervals <6 week and ≥12 weeks, and corresponding immunogenicity and efficacy, were substituted for the pooled vaccine efficacy estimate used in Figure 1. Exploratory analyses are denoted by blue dots. Ratios for neutralizing antibody titer (Panel A) and binding antibody titer (Panel B) were generated for Oxford/AstraZeneca using the HCS median titer from Phase 1/2 publication, as noted in Tables S1 and S2.

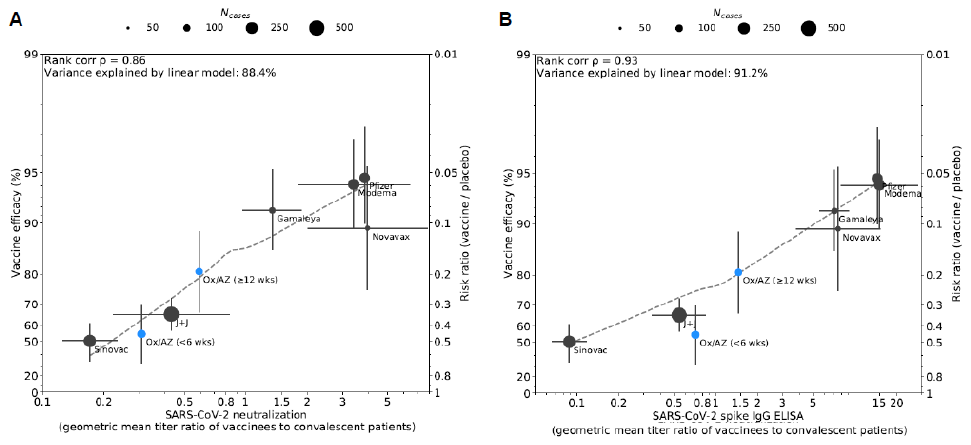

**Table S1. Data summary and sources for vaccine-induced and HCS neutralizing antibody titers.** Confidence intervals estimated for HCS panels from published figures for Oxford/AstraZeneca and Janssen/J&J. WT VNA = Wild-type virus neutralization assay. Ps VNA = Pseudovirus neutralization assay. NA = Not available.

* Denotes median titers and interquartile range, instead of geometric mean titers and 95% confidence intervals

| **Developer** | **% Efficacy (CI)** | **Cases (N)** | **Vaccine-induced** | |  | **HCS** | |  |  |  |
| --- | --- | --- | --- | --- | --- | --- | --- | --- | --- | --- |
|  |  |  | **GMT (CI)** | **N** |  | **GMT (CI)** | **N** | **Vaccine/**  **HCS ratio** | **Assay** | **HCS description** |
| **Pfizer/BioNTech** |  |  |  |  |  |  |  |  |  |  |
| Polack et al.^5^ | 94.6 (89.9, 97.3) | 178 |  |  |  |  |  |  |  |  |
| Walsh et al.^6^ |  |  | 361 (NA) | 11 |  | 94 | 38 | 3.8 | WT VNA | Asymptomatic + symptomatic (mild-severe) |
| **Moderna** |  |  |  |  |  |  |  |  |  |  |
| Baden et al.^7^ | 94.1 (89.3, 96.8) | 196 |  |  |  |  |  |  |  |  |
| Anderson et al.^8^ |  |  | 360 (273, 476) | 14 |  | 106 (60, 189) | 41 | 3.4 | Ps VNA | Symptomatic (mild-severe) |
| **Gamaleya** |  |  |  |  |  |  |  |  |  |  |
| Logunov et al. (2021)^9^ | 91.6 (85.6, 95.2) | 78 | 44.5 (32, 62) | 100 |  |  |  | 1.4 | WT VNA |  |
| Logunov et al. (2020)^10^ |  |  |  |  |  | 33 (31.5, 34.5) | 4817 |  | WT VNA | Symptomatic (mild-moderate) |
| **Oxford/AstraZeneca (Primary correlation analysis)** | | | | | | | | |  |  |
| Voysey et al.^4^ | 66.7 (57.4, 74.0) | 332 |  |  |  |  |  |  |  |  |
| Folegatti et al.^11^ |  |  | 451* (212, 628) | 9 |  | 408* (212, 2000) | 170 | 1.1 | Ps VNA | Asymptomatic + symptomatic (mild-severe) |
| **Sinovac** |  |  |  |  |  |  |  |  |  |  |
| ANVISA press briefing^12^ | 50.4 (34.1, 60.7) | 252 |  |  |  |  |  |  |  |  |
| Zhang et al.^13^ |  |  | 28 (23, 34) | 118 |  | 164 (129, 209) | 117 | 0.17 | WT VNA | Symptomatic (mild-severe) |
| **Novavax (Primary correlation analysis)** | | | |  |  |  |  |  |  |  |
| Novavax press briefing^2^ | 89.3 (75.2, 95.4) | 62 |  |  |  |  |  |  |  |  |
| Keech et al.^14^ |  |  | 3906 (2556, 5970) | 29 |  | 984 (579, 1671) | 32 | 4.0 | WT VNA | Asymptomatic + symptomatic (majority mild) |
| **Janssen/J&J (Primary correlation analysis)** | | | | |  |  |  |  |  |  |
| FDA briefing document^15^ | 65.5 (57.2, 72.4) | 437 |  |  |  |  |  |  |  |  |
| Sadoff et al.^16^ |  |  | 224 (158, 318) | 24 |  | 522 (277, 850) | 32 | 0.43 | WT VNA | Symptomatic (majority severe) |
| **Novavax (Excl. VOC, Fig. S3)** | | | | | | |  |  |  |  |
| Novavax press briefing^2^ | 95.6 (84.1, 98.5) | 56 |  |  |  |  |  |  |  |  |
| Keech et al.^14^ |  |  | 3906 (2556, 5970) | 29 |  | 984 (579, 1671) | 32 | 4.0 | WT VNA | Asymptomatic + symptomatic (majority mild) |
| **Janssen/J&J (US, Fig. S3)** | | | | | |  |  |  |  |  |
| FDA briefing document^15^ | 72.0 (58.2, 81.7) | 144 |  |  |  |  |  |  |  |  |
| Sadoff et al.^16^ |  |  | 224 (158, 318) | 24 |  | 522 (277, 850) | 32 | 0.43 | WT VNA | Symptomatic (majority severe) |
| **Oxford/AstraZeneca (< 6wks, Fig. S4)** | | | | | | | | | | |
| Voysey et al.^4^ | 54.9 (32.7, 69.7) | 111 | 125 (110, 141) | 272 |  |  |  | 0.31 | Ps VNA |  |
| Folegatti et al.^11^ |  |  |  |  |  | 408* (212, 2000) | 170 |  | Ps VNA | Asymptomatic + symptomatic (mild-severe) |
| **Oxford/AstraZeneca (≥ 12 wks, Fig. S4)** | | | | | | | | | | |
| Voysey et al.^4^ | 80.7 (66.5, 88.9) | 332 | 240 (210, 276) | 217 |  |  |  | 0.59 | Ps VNA |  |
| Folegatti et al.^11^ |  |  |  |  |  | 408* (212, 2000) | 170 |  | Ps VNA | Asymptomatic + symptomatic (mild-severe) |

**Table S2. Data summary and sources for vaccine-induced and HCS binding antibody titers.** Confidence intervals estimated from figures for HCS panel and US site titers for Janssen/J&J. Median titer estimated for HCS panel from published figure for Oxford/AstraZeneca. WT VNA = Wild-type virus neutralization assay. Ps VNA = Pseudovirus neutralization assay. NA = Not available.

* Denotes median titers and interquartile range, instead of geometric mean titers and 95% confidence intervals

| **Developer** | **% Efficacy (CI)** | **Cases (N)** | **Vaccine-induced** | |  | **HCS** | |  |  |  |
| --- | --- | --- | --- | --- | --- | --- | --- | --- | --- | --- |
|  |  |  | **GMT (CI)** | **N** |  | **GMT (CI)** | **N** | **Vaccine/**  **HCS ratio** | **ELISA antigen** | **HCS description** |
| **Pfizer/BioNTech** |  |  |  |  |  |  |  |  |  |  |
| Polack et al.^5^ | 94.6 (89.9, 97.3) | 178 |  |  |  |  |  |  |  |  |
| Walsh et al.^6^ |  |  | 9136 (NA) | 11 |  | 631 | 38 | 15 | S1 | Asymptomatic + symptomatic (mild-severe) |
| **Moderna** |  |  |  |  |  |  |  |  |  |  |
| Baden et al.^7^ | 94.1 (89.3, 96.8) | 196 |  |  |  |  |  |  |  |  |
| Anderson et al.^8^ |  |  | 558,905 (462,907; 674,810) | 14 |  | 37,244 (20,170; 68,771) | 41 | 15 | RBD | Symptomatic (mild-severe) |
| **Gamaleya** |  |  |  |  |  |  |  |  |  |  |
| Logunov et al. (2021)^9^ | 91.6 (85.6, 95.2) | 78 | 8996 (7610; 10,635) | 100 |  |  |  | 7.1 | RBD |  |
| Logunov et al. (2020)^10^ |  |  |  |  |  | 1266 (1066, 1504) | 4817 |  | RBD | Symptomatic (mild-moderate) |
| **Oxford/AstraZeneca (Primary correlation analysis)** | | | | | | | | |  |  |
| Voysey et al.^4^ | 66.7 (57.4, 74.0) | 332 |  |  |  |  |  |  |  |  |
| Folegatti et al.^11^ |  |  | 26,251* (16,453; 36,643) | 9 |  | 33,000* (NA) | 180 | 0.80 | S | Asymptomatic + symptomatic (mild-severe) |
| **Sinovac** |  |  |  |  |  |  |  |  |  |  |
| ANVISA press briefing^12^ | 50.4 (34.1, 60.7) | 252 |  |  |  |  |  |  |  |  |
| Zhang et al.^13^ |  |  | 1094 (937, 1278) | 117 |  |  |  | 0.09 | RBD | Symptomatic (mild-severe) |
| Wang et al.^1^ |  |  |  |  |  | 12,442 (9755; 15,869) | 117 |  | RBD |  |
| **Novavax (Primary correlation analysis)** | | | |  |  |  |  |  |  |  |
| Novavax press briefing^2^ | 89.3 (75.2, 95.4) | 62 |  |  |  |  |  |  |  |  |
| Keech et al.^14^ |  |  | 63,160 (47,117; 84,666) | 29 |  | 8344 (4421; 15,748) | 32 | 7.6 | S | Asymptomatic + symptomatic (majority mild) |
| **Janssen/J&J (Primary correlation analysis)** | | | | |  |  |  |  |  |  |
| FDA briefing document^15^ | 65.5 (57.2, 72.4) | 437 |  |  |  |  |  |  |  |  |
| Sadoff et al.^16^ |  |  | 478 (379, 603) | 69 |  | 879 (628, 1332) | 32 | 0.54 | S | Symptomatic (majority severe) |
| **Novavax (Excl. VOC, Fig. S3)** | | | | | | |  |  |  |  |
| Novavax press briefing^2^ | 95.6 (84.1, 98.5) | 56 |  |  |  |  |  |  |  |  |
| Keech et al.^14^ |  |  | 63,160 (47,117; 84,666) | 29 |  | 8344 (4421; 15,748) | 32 | 7.6 | S | Asymptomatic + symptomatic (majority mild) |
| **Janssen/J&J (US, Fig. S3)** | | | | | |  |  |  |  |  |
| FDA briefing document^15^ | 72.0 (58.2, 81.7) | 144 | 412 (379, 603) | 48 |  |  |  | 0.47 | S |  |
| Sadoff et al.^16^ |  |  |  |  |  | 879 (628, 1332) | 32 |  | S | Symptomatic (majority severe) |
| **Oxford/AstraZeneca (< 6wks, Fig. S4)** | | | | | | | | | | |
| Voysey et al.^4^ | 54.9 (32.7, 69.7) | 111 | 23,453 (21,040; 26,142) | 300 |  |  |  | 0.71 | S |  |
| Folegatti et al.^11^ |  |  |  |  |  | 33,000* (NA) | 180 |  | S | Asymptomatic + symptomatic (mild-severe) |
| **Oxford/AstraZeneca (≥ 12 wks, Fig. S4)** | | | | | | | | | | |
| Voysey et al.^4^ | 80.7 (66.5, 88.9) | 332 | 47,942 (43,638; 52,670) | 397 |  |  |  | 1.5 | S |  |
| Folegatti et al.^11^ |  |  |  |  |  | 33,000* (NA) | 180 |  | S | Asymptomatic + symptomatic (mild-severe) |
